# Metabolic Preference Assay for Rapid Diagnosis of Bloodstream Infections

**DOI:** 10.1101/2021.04.09.21255220

**Authors:** Thomas Rydzak, Ryan A. Groves, Ruichuan Zhang, Rajnigandha Pushpker, Maryam Mapar, Ian A. Lewis

**Affiliations:** Department of Biological Science, University of Calgary, Calgary, AB T2N 1N4, Canada

## Abstract

Bloodstream infections (BSIs) cause >500,000 infections and >80,000 deaths per year in North America. The length of time between the onset of symptoms and administration of appropriate antimicrobials is directly linked to mortality rates. It currently takes 2-5 days to identify BSI pathogens and measure their susceptibility to antimicrobials – a timeline that directly contributes to preventable deaths. To address this, we developed a rapid metabolic preference assay (MPA) that uses the pattern of metabolic fluxes observed in *ex-vivo* microbial cultures to identify common pathogens and determine their antimicrobial susceptibility profiles. In a head-to-head race with a leading platform (VITEK 2) used in diagnostic laboratories, MPA decreased testing timelines from 40 hours to under 20. If put into practice, this assay could reduce septic shock mortality and reduce the use of broad spectrum antibiotics.

**One Sentence Summary:** Metabolomics enables rapid diagnosis of BSIs.

## Main Text

The diagnostic tools used to identify pathogens and measure antimicrobial susceptibility play a critical role in controlling infectious diseases. In the case of bloodstream infections (BSIs), rapid diagnostic timelines are critical because a patient’s odds of surviving an infection are inversely proportional to the length of time that elapses between the onset of symptoms and the administration of appropriate antimicrobials^1,2^. A single day of receiving an ineffective antimicrobial increases average BSI mortality by up to 5%^3,4^. When BSIs progress into septic shock, these treatment delays can increase mortality by over 7% per hour (Fig. 1)^2^.

**Fig. 1.**
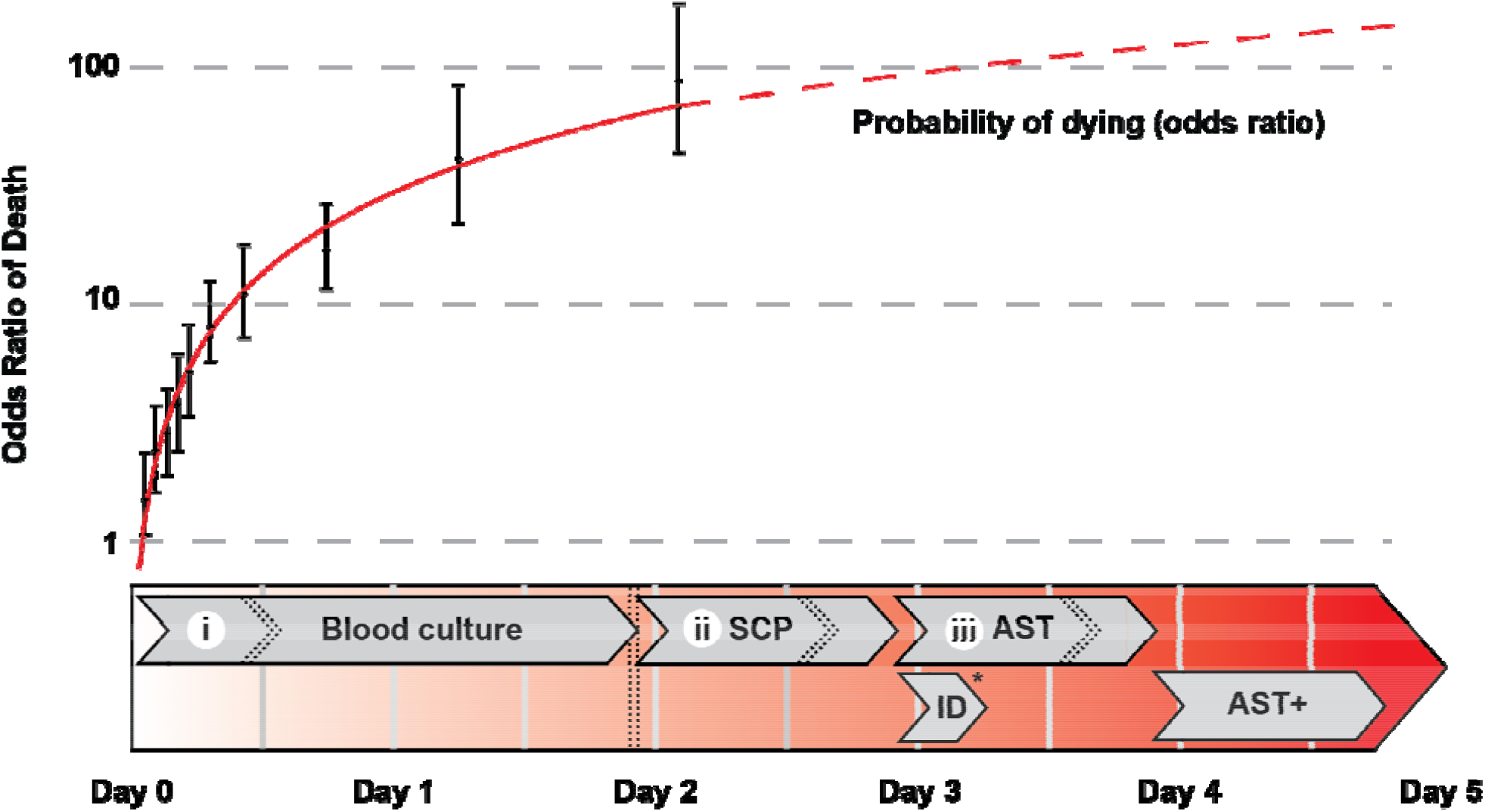
BSI testing timelines versus mortality rates. Mortality risk (expressed as adjusted odds ratio of death) adapted with permission from Kumar et al., 2006 shown between the onset of symptoms and administration of antimicrobials (top) relative to current clinical testing times (bottom). Error bars represent the 95% confidence interval of the odds ratio. ID, microbial identification by MALDI-TOF-MS; SCP, single colony purification on agar plates; AST, antimicrobial susceptibility testing; AST+, additional antimicrobial susceptibility testing for isolates with an unusual resistance profile.Roman numerals represent the rate limiting microbial culturing steps. Dashed lines show the median culture timelines observed in high-volume diagnostic laboratories whereas the solid lines show the range of analysis times for each step. *While ID can be performed either after SCP or directly from blood culture bottles, the need for SCP for current AST methods means that overall timelines are unaffected.

Unfortunately, most diagnostic laboratories require 2-5 days to complete microbial identification (ID) and antimicrobial susceptibility testing (AST; Fig. 1). Currently, 20-30% of patients are prescribed the wrong antimicrobial treatment^5,6^, a percentage that is expected to rise with the increasing prevalence of antimicrobial resistant organisms. Given that 1.3 million people suffer from BSIs in North America and Europe each year, and roughly 182,000 of these people die^7^, a more efficient method for completing ID and AST could save tens of thousands of lives each year, decrease in-patient hospital stays, and decrease average treatment costs^8,9^. In addition, long diagnostic times contribute to the selection of antimicrobial resistant organisms: long testing timelines combined with the time-sensitive nature of treating BSIs creates a circumstance in which clinicians must make therapeutic decisions with little or no laboratory data. This situation encourages the widespread use of broad-spectrum antimicrobials and selection for resistant organisms^10^. In summary, any new technology that shortens the diagnostic timeline will have a significant impact on global health^1,11,12^.

### Factors limiting BSI diagnostic timelines

The current clinical microbiology testing timeline is dominated by three microbial culturing steps that are necessary for identifying and characterizing bloodstream pathogens (Fig. 1). These microbial amplification steps are the rate-limiting factors in the analysis pipeline and are the primary technical roadblocks to faster diagnostics. In the first culturing step *(i)* patient blood samples are incubated for 7-43 h^13^ to allow microbes present in the samples to grow to detectable densities (from 0.01-100 CFU/mL to >1×10^9^ CFU/mL^14,15^). Although the median time to positivity is 12.7 h, this step can require as much as 125 h for some slow growing organisms^16,17^. Once blood cultures have flagged positive, *(ii)* aliquots of cultures are streaked onto agar plates and incubated for another 12-24 h^18^ to obtain single colonies. These sub-cultured isolates are then identified via matrix-assisted laser desorption/ionization time-of-flight (MALDI-TOF) mass spectrometry (MS)^19^. Antimicrobial susceptibility testing is then performed by *(iii)* incubating a fixed number of microbes (5×10^5^ CFU/mL) for 12-24 h in a medium containing antimicrobials using an automated testing system (e.g., VITEK 2, BioMérieux; MicroScan, Beckman Coulter; Sensititre, Thermo Fisher Scientific; Phoenix, BD)^20^. Additional antimicrobial testing procedures may also be necessary for isolates with an unusual resistance profile^21^. In summary, current clinical diagnostic timelines are limited by microbial growth rates.

Both cost constraints and health concerns have created considerable pressure to develop a faster clinical diagnostic pipeline ^22^. Some time savings have been achieved by streamlining the existing workflow. Direct MALDI-TOF-MS analysis of blood cultures, for example, allows microbes to be identified faster by circumventing one microbial culture step (Fig. 1 ***ii***)^23^. Unfortunately, direct MALDI-TOF-MS cannot replace the existing AST workflow. An alternative emerging strategy has been to use DNA-based technologies to both identify organisms and detect common resistance genes in a single multiplexed assay (e.g., Biofire® FilmArray® and Verigene® ^24^). Though promising, these assays have limitations: they require culture-based isolation of the pathogen, are susceptible to false-negative results due to primer specificity or PCR inhibition, and can give false positive results due to cell-free DNA^25,26^. Moreover, both proteomic and DNA-based assays detect the genetic potential for drug resistance, not the empirically-determined antimicrobial susceptibility phenotype^27^. Any genetic modulators of resistance, or novel resistance mechanisms, cannot be detected via these assays. Consequently, direct phenotypic assessment of susceptibility via microbial culturing is required by the current clinical laboratory guidelines^21^.

### Advantages of metabolomics-based platforms for BSI diagnostics

Although faster molecular-based diagnostic tools are emerging, the need to both identify organisms and empirically determine their antimicrobial resistance profiles has prevented many of these tools from being integrated into working diagnostic laboratories. Metabolomics offers a unique opportunity to accelerate diagnostics while conforming to the established workflow used in clinical practices. Metabolites are thousands of times more abundant than individual proteins^28^, are sensitive reporters of microbial physiology^29^, and are compatible with established high-throughput clinical mass spectrometry platforms^30^. As a result, sensitive metabolite-based assays have the potential to minimize the rate-limiting steps in the existing clinical workflow. Although the applicability of metabolomics to microbial diagnostics has been recognized for over a decade^31-36^, previous applications have largely sought to identify native biomarkers present in blood of people with infections. This approach is challenging due to the intrinsic variability of human metabolism. Moreover, it does not provide a mechanism for assessing antimicrobial susceptibility.

Herein, we introduce a new diagnostic strategy, the metabolic preference assay (MPA), that uses the patterns of consumed versus excreted metabolites of *ex-vivo* microbial cultures to both identify pathogens and measure their antimicrobial susceptibility. We identify biomarkers capable of differentiating seven of the most prevalent organisms responsible for BSIs [*Candida albicans* (CA), *Klebsiella pneumoniae* (KP), *Escherichia coli* (EC), *Pseudomonas aeruginosa* (PA), *Staphylococcus aureus* (SA), *Enterococcus faecalis* (EF), and *Streptococcus pneumoniae* (SP)]^37^, show that changes in metabolite levels can empirically measure activity of antimicrobials, and demonstrate that the MPA can produce results in a fraction of the time as current testing methods.

### Biomarkers can differentiate between prevalent species responsible for of BSIs

Although the core architecture of central carbon metabolism is shared among almost all organisms, the activities of these pathways differ according to both genetic and environmental factors. Thus, by tightly controlling environmental variables, metabolic pathway activities should serve as indicators of microbial species. This fundamental hypothesis is the root of the MPA-based diagnostic approach and is testable by quantifying metabolic boundary fluxes (the rates at which nutrients and waste products are consumed or produced) of clinically-relevant microbes under well-controlled conditions. To test this, the metabolic boundary fluxes of the 7 most common bloodstream pathogens were measured. For initial biomarker discovery, three clinical isolates (2 for PA) from each target species (n = 7) were analyzed in replicate (n = 9). Microbial cultures were seeded at a 0.5 McFarland (OD_600_ ∼ 0.07 or 1.5 × 10^8^ CFU/mL) into Mueller Hinton broth containing 10% human blood (MHB). Metabolite levels present in the cultures were analyzed at 0 h and 4 h on a Thermo Q Exactive HF MS in negative mode. Untargeted analysis identified 799 peaks with significant fold changes between species (as computed by analysis of variance using a Bonferroni corrected α = 0.05; Extended Data File 1; Extended Data File 2). These signals were clustered into 104 groups on the basis of retention times, known adduct/fragment masses, and covariance of signal intensities across replicates using R software developed in house. The most likely parent ion for each biomarker was then identified from each of the 104 groups based on signal intensities (Extended Data File 3). Putative metabolite assignments were made via the Madison Metabolomics Consortium Database (MMCD)^38^ and Human Metabolome Database^39^, and select metabolites were confirmed via MS/MS fragmentation and by standard addition (Extended Data Fig. 1; Extended Data File 4).

Species-dependent consumption or production of our 104 biomarkers robustly differentiates between the seven target species (Fig. 2A). Although the overall pattern of markers was similar between closely related microbes (i.e. *K. pneumoniae* and *E. coli*), they could still be differentiated via select biomarkers. Remarkably, just seven production biomarkers were sufficient to distinguish between the target pathogens and acted as binary predictors of each species (Fig. 2B). Specifically, arabitol, xanthine, and N^1^,N^12^-diacetylspermine were exclusively produced by *C. albicans, P. aeruginosa*, and *E. faecalis*, respectively. Both *K. pneumoniae* and *E. coli* produced succinate, but the latter did not produce urocanate. Mevalonate was produced by *S. aureus*, and to a lesser extent *E. faecalis*, but unlike *E. faecalis, S. aureus* did not produce N^1^,N^12^-diacetylspermine. Lactate was produced by *S. pneumoniae*, and to a lesser extent, *E. faecalis*. Notably, addition of 10% blood to the medium, irrespective of donor (n = 20), had negligible effects on metabolite profiles when compared to seeded culture, demonstrating that these biomarkers were pathogen-specific, and did not reflect blood metabolism or donor-specific metabolite carryover (Extended Data Fig. 2; Extended Data File 5). With the exception of arabitol production by *C. albicans*^40^ and fermentative succinate production by *E. coli* and *K. pneumoniae*^41,42^, secretion of the species-specific biomarkers mentioned above have not been previously identified. These data show that the changing patterns of metabolites present in microbial cultures can be harnessed as a robust diagnostic tool for identifying bloodstream pathogens.

**Fig. 2.**
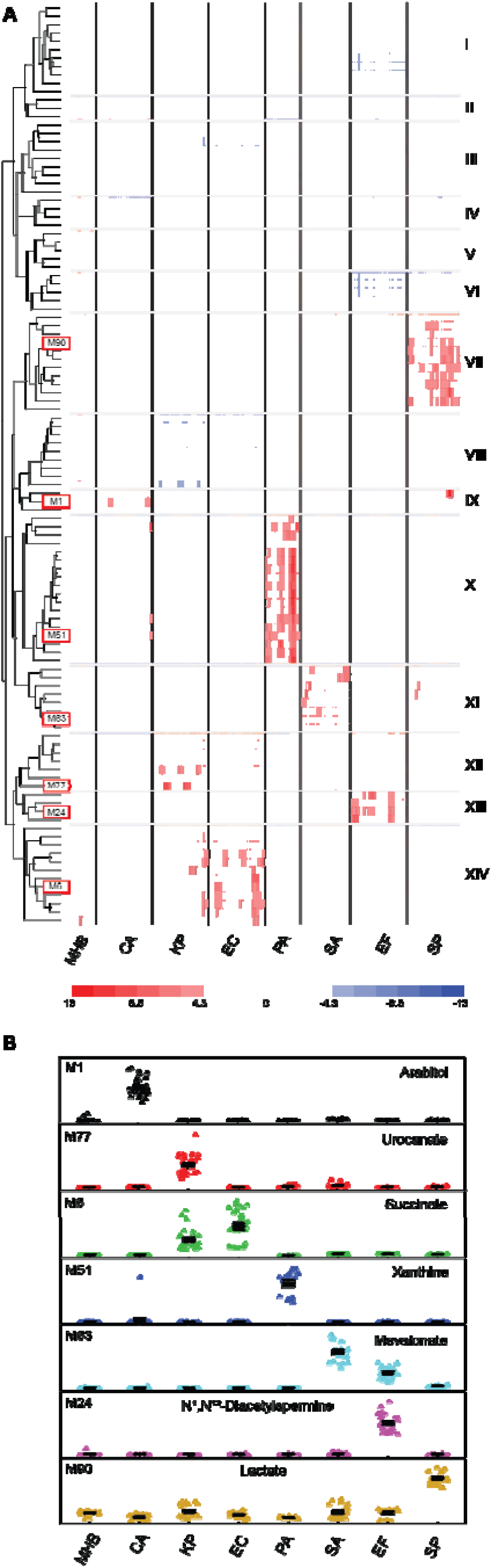
Metabolic preference assay of seven pathogens responsible for 85% of bloodstream infections. (**A**) Heat map of the top 104 biomarkers after a 4 h incubation period. Biomarkers were selected from 4,372 signals observed in LC-MS spectra of the discovery dataset (3 isolates of each species (2 for PA) performed with 9 replicates; Extended Data Files 1-3). Biomarkers were clustered into groups based on co-retention, m/z shift relative to known fragments/adducts, and co-variance. Marker numbers (M) in (A) correspond to metabolites in (B). (**B**) Top seven biomarkers that can robustly differentiate between the seven species studied. MHB; Mueller Hinton broth with 10% blood.

### Rapid antibiotic susceptibility testing by MPA

One major advantage of using metabolomics for microbial diagnostics is that metabolism is a sensitive reporter of cell physiology. Nutritional precursors are converted into waste products at rates that are many orders of magnitude faster than microbial growth. Moreover, these processes are dramatically altered, or halted completely, when cells are exposed to toxic substances. Consequently, metabolomics approaches offer a unique opportunity to empirically assess antibiotic sensitivity in a fraction of the time that is required by current growth-based AST approaches. Herein, we evaluate the practicality of using a MPA-based metabolic inhibition assay (MIA) as a diagnostic platform for quantifying AST.

MIA was accomplished by monitoring changes in the metabolic composition of microbial culture supernatants after a 4 h incubation period with and without antimicrobials. Microbes were seeded into MHB medium at 10% of a 0.5 McFarland (to a final OD_600_ of ∼0.007) and metabolomics analyses were conducted using the same method used for general MPA testing. The metabolic concept underpinning MIA is illustrated in the top left panel of Fig. 3: drug sensitive strains of *K. pneumoniae* show an incremental reduction in hypoxanthine production proportional to meropenem concentrations whereas resistant strains remain unaffected within the clinically-relevant antibiotic concentrations. Similar antibiotic-induced metabolic perturbations were observed in all of the target pathogens when isolates were exposed to minimum inhibitory concentrations of commonly prescribed antimicrobials (Fig 3).

**Fig. 3.**
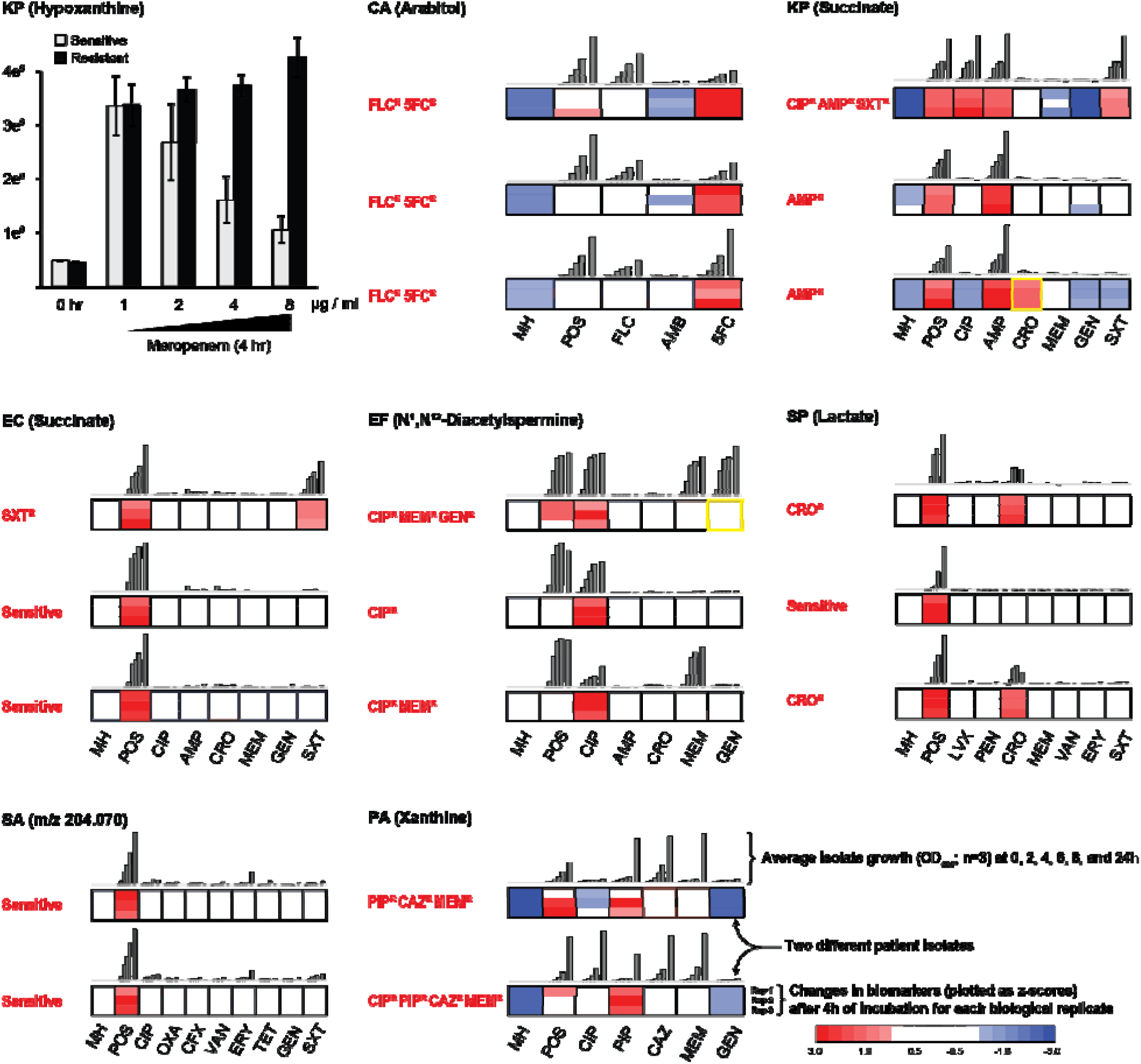
Metabolic inhibition assay (MIA) for assessing antimicrobial susceptibility. Concentrations of select biomarkers produced by a representative cohort of clinical isolates (7 species, 2-3 isolates each, performed in triplicate) were quantified after a 4 h incubation in the presence of antimicrobials. The top-left panel demonstrates the MIA concept where a concentration-dependent decrease in hypoxanthine production in response to meropenem dose is observed, an effect that is not observed in drug sensitive isolates. Remaining panels show the metabolic responses of pathogens exposed to antimicrobials prepared at the minimum inhibitory CLSI breakpoints differentiating sensitive versus intermediate resistant isolates (*21*). The empirically-determined susceptibility profiles, as measured by VITEK 2, are reported in red text alongside each panel, and were confirmed independently by growth assays (grey bars). All MIA-determined susceptibility profiles matched those reported by the VITEK 2 with only two exceptions (highlighted in yellow boxes), resulting in a 2% false discovery rate for the assay (1% very major error and 1% major error by CLSI guidelines). Abbreviations (MIC in µg/mL): FLC, fluconozole (2); AMB, amphotericin B (2); 5FC, 5-flucytosine (0.5); CRO, Ceftriaxone (0.5 and 1 for *S. pneumoniae* and *K. pneumoniae*/*E. coli* respectively); CIP, ciprofloxacin (1); MEM, meropenem (0.25, 1, and 2 for *S. pneumoniae, K. pneumoniae*/*E. coli*, and *P. aeruginosa*, respectively); GEN, gentamicin (4; 500 only for *E. faecalis*); AMP, ampicillin (8); SXT, trimethoprim-sulfamethoxazole (2/38); CAZ, ceftazidime (8); PIP, piperacillin (16); LVX, levofloxacin (2); PEN, penicillin (0.06); ERY, erythromycin (0.25 and 0.5 for *S. pneumoniae* and *S. aureus*, respectively); OXA, oxacillin (2); VAN, vancomycin (1, 2 and 4 for *S. pneumoniae, S. aureus*, and *E. faecalis*, respectively); CFX, cefazolin (4); TET, tetracycline (4).

To assess MIA as a potential clinical tool, three patient isolates for each target pathogen (2 for *S. aureus* and *P. aeruginosa*) were analyzed. Antifungals (azoles, polyenes, and antimetabolites) were tested for *C. albicans*. Both bactericidal (penicillins, cephalosporins, carbapenems, glycopeptides, aminoglycosides, fluoroquinolones and trimethoprim-sulfamethoxazole) and bacteriostatic (macrolides and tetracyclines) antibiotic classes were evaluated. Antimicrobial sensitivity profiles determined by MIA were consistent with 98% of the profiles observed in traditional microbial growth assays. The assays were consistent across all antimicrobial’s mechanisms of action (Fig. 3; Extended Data File 6). For example, succinate production by ampicillin (AMP) and trimethoprim/sulfamethoxazole (SXT) resistant *E. coli* was comparable when the strain was incubated in the presence of AMP, SXT, or in the absence of antibiotics. However, succinate production was significantly lower (p < 0.01 for all pairwise comparisons) when the strain was grown in the presence of antibiotics to which it was sensitive. In most cases, the biomarkers used to identify microbes were also useful for differentiating drug sensitive and resistant strains (e.g., arabitol for *C. albicans*, succinate for *K. pneumoniae* and *E. coli*, N^1^,N^12^-diacetylspermine for *E. faecalis*, xanthine for *P. aeruginosa*, and lactate for *S. pneumoniae*). One exception to this trend was mevalonate, which is an excellent marker for *S. aureus* but an unreliable marker for drug resistance. Instead, an alternative compound with an *m/z* of 204.069 was identified as a more stable metric for differentiating resistant versus susceptible strains of *S. aureus*. Collectively, these data indicate that the MIA strategy can be a robust indicator of antimicrobial resistant profiles.

### Metabolic-based diagnostics decreases pathogen ID and AST timelines by more than 20 hours

One of the primary motivations for this project is the urgent need for rapid diagnostic tools for bloodstream infections. To evaluate the potential time savings available via our MPA/MIA diagnostic workflow, we conducted three independent head-to-head races between the MPA and the bioMéieux VITEK 2 platform. Aerobic BacT/Alert bottles were seeded with 10 mL of blood and 1 mL of diluted culture containing 40-60 CFU/mL of exponential phase bacteria (*S. aureus* and *E. coli*, n = 3 each), and were incubated in a BacT/Alert 3D automated microbial detection system (bioMérieux) until the bottles flagged positive. One aliquot from each bottle was taken for testing on the VITEK 2, which involved plating an aliquot on blood agar plates, incubating for 18 hours, picking colonies, diluting cultures according to the manufactures protocols, and loading samples onto VITEK ID and AST cards. Notably, ID and AST was performed simultaneously on the VITEK. A second aliquot from each bottle was used for identifying and characterizing pathogens via the MPA/MIA workflow. For MPA/MIA analyses, media containing the most commonly tested antibiotics for each strain (CFZ, OXA, AMP, SXT, and CIP for *S. aureus*; AMP, GEN, SXT and CIP for *E. coli*) at concentration ranges consistent with MicroScan Panels were inoculated with 0.5% of the positive blood-bacterium-BacT/Alert medium mixture. Samples were processed following the 4 h incubation period and analyzed via LC-MS using a 5 min HILIC method. Data were analyzed in real-time using the MAVEN software package^43^. A positive growth control (medium with no antibiotic) was analyzed first to enable species identification. Subsequently, only samples incubated in the presence of antibiotics pertinent to the identified species were analyzed in order to minimize MS analysis time. Our MIA results agreed with CLSI MIC breakpoints validating that the process can be performed directly from BacT bottles. The MPA/MIA workflow reduced total testing time by an average of 24.3 h for *S. aureus* and 22.4 h for *E. coli*, corresponding to a 2.2 and 2.3-fold decrease in total testing time, respectively (Fig. 4). The time required for strain identification (*ii*) and antibiotic susceptibility (*iii*) alone decreased by 4.7 and 5.0-fold for *S. aureus* and *E. coli*, respectively.

**Fig. 4.**
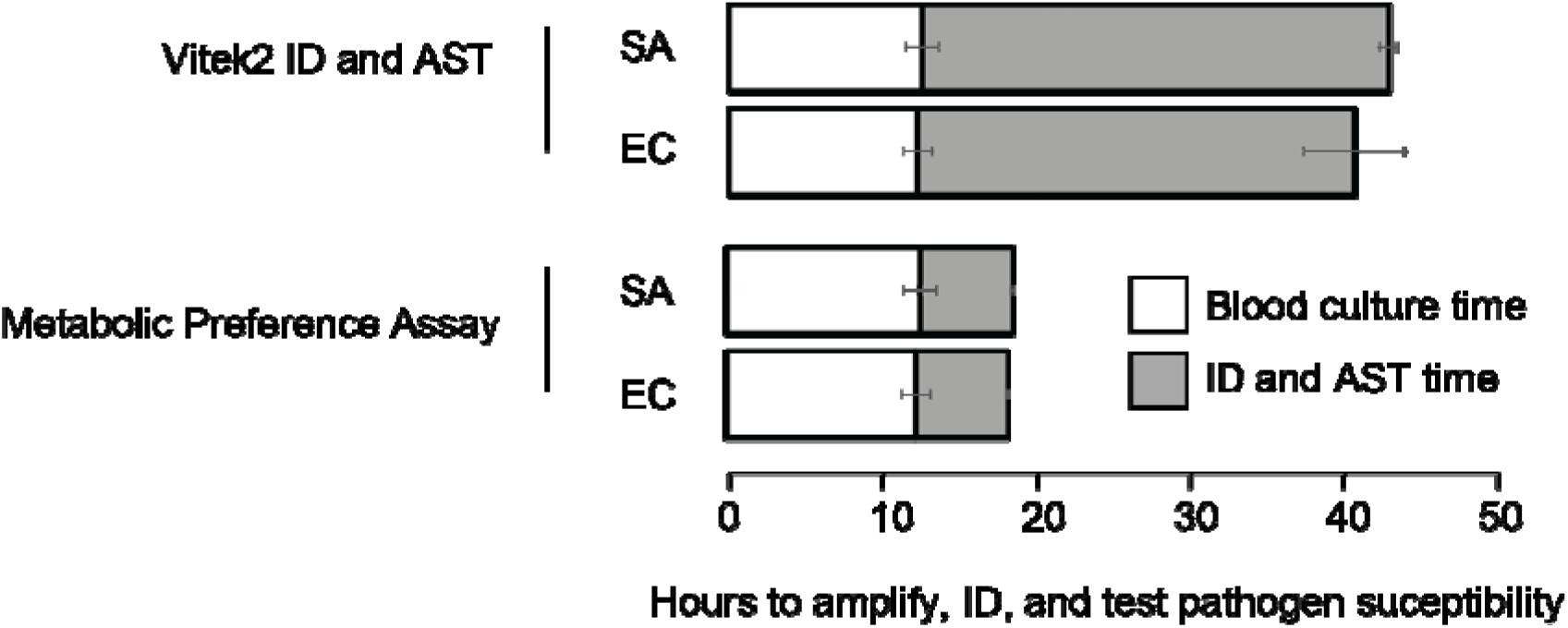
Diagnostic timelines from a head-to-head race between the VITEK 2 testing platform versus our metabolomics-based workflow. Total time to identify and perform AST using our MPA/MIA metabolomics workflow was more than 2.2-fold faster than current clinical methods, whereas identification and testing times alone were more than 4.7-fold faster. The observed 22 h decrease in testing time allows for administration of correct antimicrobial therapies, which could reduce mortality rates in septic shock patients.

## Discussion

Rapid diagnostic tools for identifying bloodstream infections could significantly improve survival rates and could help reduce the widespread use of broad spectrum antimicrobials. The culture-based diagnostic workflow (Fig. 1) is currently the ‘gold standard’ process for identifying bloodstream pathogens and measuring their antimicrobial breakpoints in most clinical microbiology laboratories. Unfortunately, this established process takes two to five days to complete, which directly contributes to negative outcomes from these serious infections. To address the urgent need for faster diagnostics, we have developed a novel metabolomics-based strategy for identifying bloodstream pathogens and empirically measuring their antimicrobial susceptibilities. Our approach provides comparable clinical data in a fraction of the time — >20 hours faster than the current practice.

The importance of rapid BSI testing is widely recognized and has driven the development of diverse molecular, chemical, and optical tests as alternatives the current pipeline^44-50^. Although many of these emerging technologies have shown promise, each new platform has limitations that prevent them from replacing the existing workflow. One of the most successful recent advancements has been the introduction of MALDI-TOF, which has enabled rapid identification of microbes. Though powerful as an identification tool, this technology does not measure antibiotic susceptibility profiles and thus must be paired with time-consuming conventional AST workflows. Similarly, multiplexed PCR and other DNA-based assays have emerged as new diagnostic alternatives to the traditional clinical microbiology workflow. These assays are attractive because they can both identify pathogens and screen for a limited number of known resistance genes. Unfortunately, the presence of resistant alleles is not a direct reporter of susceptibility. Polygenic interactions, changes in expression, novel antimicrobial efflux pumps, point mutations altering primer binding, polymorphisms modulating resistance profiles, and novel resistance mechanism can have a profound impact on a microbe’s empirically-determined antimicrobial susceptibility. Furthermore, neither MALDI-TOF nor DNA-based assays can determine antimicrobial breakpoints, which are critical for guiding prescribing practice. Consequently both MALDI and DNA-based analyses must be used in conjunction with traditional AST, which is one of the main rate limiting steps in the speed of diagnostics.

In contrast, the metabolomics workflow we introduce here delivers both microbial identities and empirically-determined susceptibility profiles that align with the requirements of current clinical practice. Moreover, the high sensitivity of mass spectrometry, along with the high abundance of metabolites relative to cells or macromolecules, make the MPA/MIA workflow inherently amenable to rapid diagnostics. Lastly, the MPA/MIA workflow can be implemented on existing clinical liquid chromatography mass spectrometers and supports the high sample throughputs needed in real-world clinical laboratories.

In summary, we demonstrate that a novel *ex-vivo* metabolomic workflow can (i) accurately identify the most common bloodstream pathogens, (ii) identify new pathogens on-the-fly by analyzing the pattern of microbial metabolites, (iii) empirically determine antibiotic susceptibility profiles, and (iv) decrease diagnostic testing timelines of bloodstream infection by more than 20 hours.

## Materials and Methods

### Experimental design

The metabolic preference assay (MPA), which measures supernatant biomarker production and consumption, was used to differentiate between 7 different species (*Candida albicans, Klebsiella pneumoniae, Escherichia coli, Pseudomonas aeruginosa, Staphylococcus aureus, Enterococcus faecalis*, and *Streptococcus pneumoniae*) responsible for 85% of bloodstream infections. Strains were grown in Mueller Hinton medium for 4 hours, and supernatants were analyzed by ultra-high performance liquid chromatography (UHPLC) MS to identify potential biomarkers. For initial biomarker discovery, three clinical isolates of each species (2 for PA) were each analyzed in replicate (n = 9). Untargeted MS analysis was used to find biomarkers that were common to all three isolates of each species, and that could differentiate between the different species. To test the reliability of MPA for antibiotic susceptibility testing (AST), changes in metabolite concentrations were measured for each of the original strains used for biomarker discovery (n=3; n=2 for *S. au* and *P. ae*) grown in the presence of antibiotics in triplicate. The metabolic inhibition assay (MIA) was further evaluated over a larger isolate cohort (n = 273) and reliable markers and threshold levels of susceptible isolates were refined. Lastly, to evaluate time savings of MPA over current state-of-the art clinical methods, a real-time head-to-head identification (ID) and AST race was performed against the VITEK 2 platform using *E. coli* and *S. aureus* on three separate occasions (see below).

### Strains, growth, and sample preparation

Isolates used in this study were recovered from patient blood culture samples and provided as cryo stocks by Alberta Precision Laboratories. Patient blood samples were also provided by (APL). The protocol was approved by the University of Calgary’s Conjoint Health Research Ethics Board under certificates # REB17-1525. All chemicals were obtained from Sigma-Aldrich (St. Louis, Mo. USA), VWR (Radnor, Pa. USA), or Fisher Scientific (Waltham, Mass. USA) unless otherwise specified. With the exception of *S. pneumoniae*, all strains were routinely grown in Mueller Hinton medium (BD Difco, Mississauga, ON, Canada). *S. pneumoniae* isolates were first revived on trypticase soy agar plates containing sheep blood (BD BBL, Mississauga, ON, Canada) and then sub-cultured into Mueller Hinton medium supplemented with catalase (1000 U/mL). For biomarker discovery using the MPA, exponential phase cultures were used to seed 96 well culture plates (Corning, New York, N.Y. USA) containing Mueller Hinton medium with 10% donated human blood to a 0.5 McFarland (OD_600_ ∼ 0.07 or ∼1.5 × 10^8^ CFU/mL). Cultures were incubated in a humidified incubator (Heracell VIOS 250i Tri-Gas Incubator, Thermo Scientific, Waltham, Mass. USA) under a 5% CO_2_ and 21% O_2_ atmosphere for four hours. After incubation, samples were transferred to a 96 well PCR plate (VWR), and centrifuged for 10 minutes for 4000 g at 4°C to remove cells. Supernatant was removed, mixed 1:1 with 100% LC-MS grade methanol, and either frozen at −80°C for further processing, or centrifuged again for 10 minutes at 4000 g at 4°C to remove any protein precipitate. Supernatant was then diluted 1:10 with 50% LC-MS grade methanol and analyzed using UHPLC-MS. AST MPAs were performed as described above, however, cultures were seeded at a 0.05 McFarland, and no blood was added to allow for periodic growth measurements at OD_600_ (Mutiskan GO, Thermo Fisher Scientific, Waltham, Mass. USA). Antibiotics used for each species was based on prevalence of being used for treatment. Published strain-specific minimum inhibitory concentrations (MIC) of each antibiotic were used^21^.

### UHPLC-MS

All metabolomics data were acquired at the Calgary Metabolomics Research Facility (CMRF). Metabolite samples were resolved via a Thermo Fisher Scientific Vanquish UHPLC platform using hydrophilic interaction liquid chromatography (HILIC). Chromatographic separation was attained using a binary solvent mixture of 20 mM ammonium formate at pH 3.0 in LC-MS grade water (Solvent A) and 0.1% formic acid (% v/v) in LC-MS grade acetonitrile (Solvent B) in conjunction with a 100 mm × 2.1 mm Syncronis™ HILIC LC column (Thermo Fisher Scientific) with a 2.1µm particle size. For general metabolic profiling runs (15 minute) the following gradient was used: 0-2 min, 100 %B; 2-7 min, 100-80 %B; 7-10 min, 80-5 %B; 10-12 min, 5% B; 12-13 min, 5-100 %B; 13-15 min, 100 %B. For expedited runs (5 minute) used for evaluation of MIA over a large cohort and race experiments, the gradient was as follows: 0-0.5 min, 100 %B; 0.5-1.75 min, 100-80 %B; 1.75-3 min, 80-5 %B; 3-3.5 min, 5% B; 3.5-4 min, 5-100 %B; 4-5 min, 100%B. The flow rate used in all analyses was 600 uL/min and the sample injection volume was 2 uL. Samples were ionized by electrospray using the following conditions: spray voltage of −2000 V, sheath gas of 35 (arbitrary units), auxiliary gas of 15 (arbitrary units), sweep gas of 2 (arbitrary units), capillary temperature of 275°C, auxiliary gas temperature of 300°C. Positive mode source conditions were the same except for the spray voltage being +3000 V. Data were acquired on a Thermo Scientific Q Exactive™ HF (Thermo Scientific) mass spectrometer using full scan acquisitions (50-750 m/z) with a 240,000 resolving power, an automatic gain control target of 3e^6^, and a maximum injection time of 200 ms. All data were acquired in negative mode except for MS/MS fragmentation analysis and confirmation of N^1^,N^12^-diacetylspermine, which ionized more efficiently in positive mode. Select biomarkers were confirmed using MS/MS analysis across a range of collision energies from 10-50 eV, at 30,000 resolving power, with a 5e^4^ automatic gain control target, and an isolation window of 4 m/z, selecting for previously observed parent ions. Biomarkers were matched to standards using fragmentation spectra and retention times. N^1^,N^12^-diacetylspermine was purchased from Cayman Chemical Company (Ann Arbor, Mich. USA), all other standards were purchased from Sigma-Aldrich. Fragmentation data were analyzed using Xcalibur 4.0.27.19 software (Thermo Scientific). All other MS analyses were conducted in MAVEN^43^.

### Real-Time ID and AST Race

*E. coli* and *S. aureus* were first grown overnight on tryptic soy agar plates and diluted in saline solution to ∼40-60 CFU/mL. Aerobic BacT/Alert bottles were seeded with 10 mL of blood and 1 mL of diluted culture containing 40-60 CFU/mL of exponential phase bacteria (*S. aureus* and *E. coli*, n = 3 each), resulting in a final cell concentration of 1-1.5 CFU/mL bottle. Bottles were immediately incubated in the BacT/Alert® automated blood culture microbial detection system. Once bottles flagged, one aliquot was used for the VITEK 2 testing pipeline (see above), and a second aliquot was used for MPA. Medium containing the most commonly tested antibiotics for each strain (CIP, OXA, SXT, CFZ, and AMP for *S. aureus*; CIP, SXT, AMP, and GEN for *E. coli*) at concentration ranges consistent with the VITEK 2 automated system were inoculated with 0.5% of the positive blood-bacterium-BacT/Alert medium mixture. Samples were processed following the 4 h incubation period and analyzed via UHPLC-MS using a 5 minute HILIC-MS method. Data was analyzed on the fly using the MAVEN software packages. The positive control (medium with no antibiotic) was analyzed first to enable species identification. Subsequently, only samples incubated in the presence of antibiotics pertinent to the identified species were analyzed in order to minimize MS analysis time.

### Statistical Analysis

Untargeted biomarkers in the preliminary dataset (7 species, 3 isolates, 9 replicates) were identified by peak picking the data in Maven with a 10 ppm m/z window and a minimum peak intensity set to 50,000. All subsequent analyzes were conducted using the R statistical software platform^52^ using in-house software tools. Untargeted analysis identified 4,372 signals in the mass spectra (Extended Data File 1). This list was ranked according to p value (as computed by analysis of variance) and 1,758 signals were identified as significant in the 4 h time point after Bonferroni correction (α = 0.05; Extended Data File 2). These signals were further thresholded (peak area-top >20,000) and fold change (pairwise mean difference between species >2 fold) resulting in 799 peaks. These signals were then clustered into 104 groups using a weighed probability function accounting for retention times, common adduct/fragment/isotopomer masses, and covariance of signal intensities among all replicates (equally weighted) using custom R software. The most likely parent ion was then selected from each cluster on the basis of signal intensity and evaluated by manual inspection of the original MS data (Extended Data File 3). Parent ions for each biomarker were then assigned using a combination of informatics tools (Madison Metabolomics Consortium Database^38^, and the Human Metabolome Database^39^). Putative metabolite assignments were then validated by purchasing standards and conducting MS/MS fragmentation and standard addition experiments (Extended Data File 4).

## Data Availability

Data is included in supplemental files.

## Funding

This work was supported by Genome Canada (10019200), Genome Alberta (10021232), Canadian Institute of Health Research (10020019), Calgary Laboratory Services (RS-16802), and the University of Calgary (Biomedical Engineering 10011121). I.A.L and R.P. are supported by an Alberta Innovates Translational Health Chair (10010625). T.R. is supported by an Eyes High Postdoctoral Fellowship from the University of Calgary (10011121). Metabolomics data were acquired by R.A.G. and R.Z. at the Calgary Metabolomics Research Facility, which is supported by the International Microbiome Centre and the Canada Foundation for Innovation (CFI-JELF 34986).

## Author contributions

T.R., R.A.G., and I.A.L. designed the experiments and analyzed the data. T.R., R.A.G. and R.Z. collected and interpreted mass spectrometry data. T.R. and R.P. conducted the microbial growth and diagnostic experiments. T.R., and I.A.L. wrote the manuscript.

## Competing interests

Drs Lewis and Rydzak are authors on a patent relating to the use of LC-MS for detecting blood stream infections. No other competing interests declared.

## Data and materials availability

All data is available in the main text or the extended data files.

## Ethics

This study was approved by the conjoint health research ethics board (REB 17-1524).

## Acknowledgements

We would like to thank APL staff for providing strains and support.

## Extended Data Materials for

**Extended Data Fig. 1.**
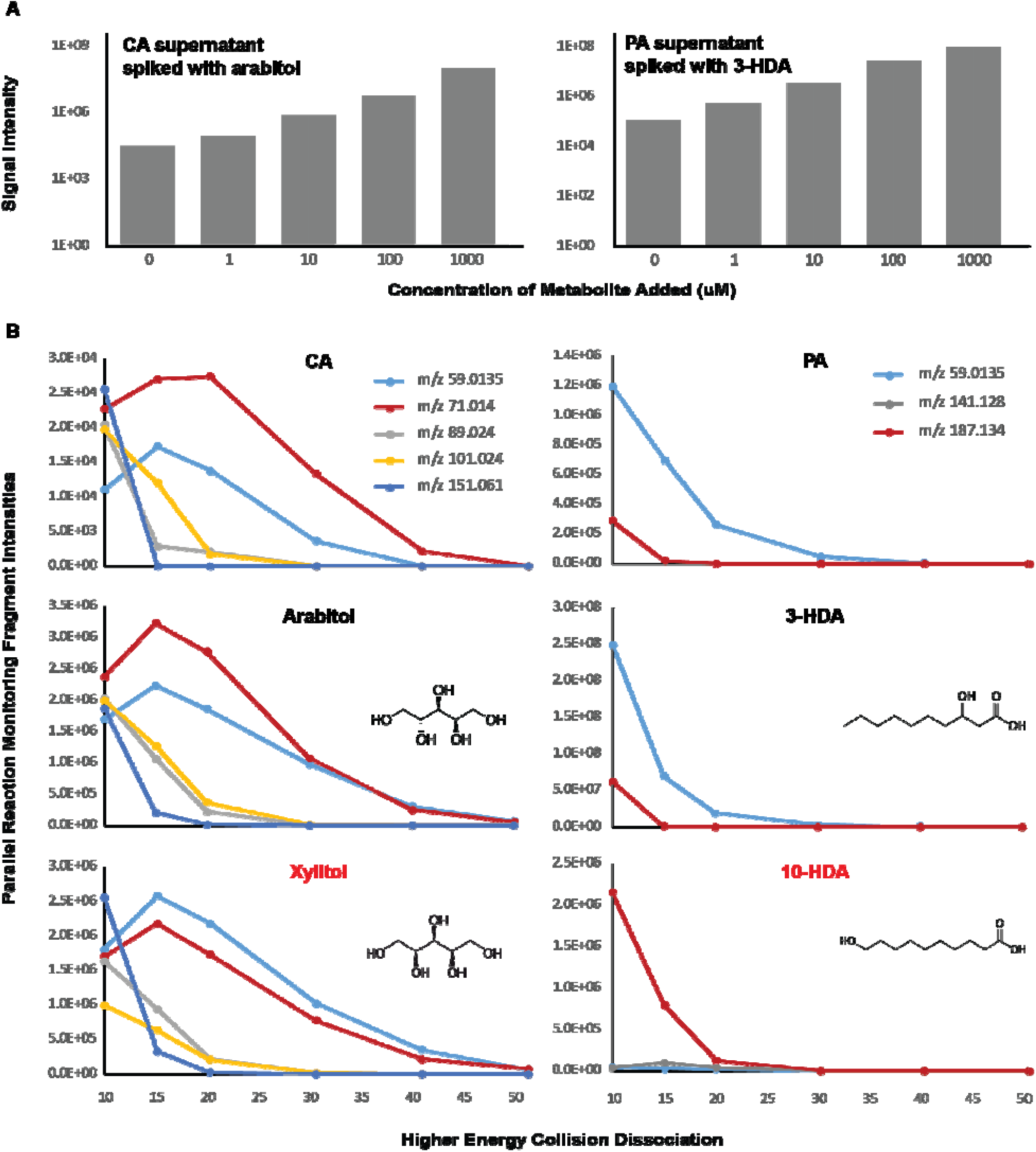
Examples of standard addition and MS/MS fragmentation patterns used to identify biomarkers. Standard additions were used to validate correct retention times of samples. In cases where the MS signal could arise from different isomers with similar retention times, MS/MS was used to identify the correct isomer. All standard addition and PRM data can be found in Extended Data File 4.

**Extended Data Fig. 2.**
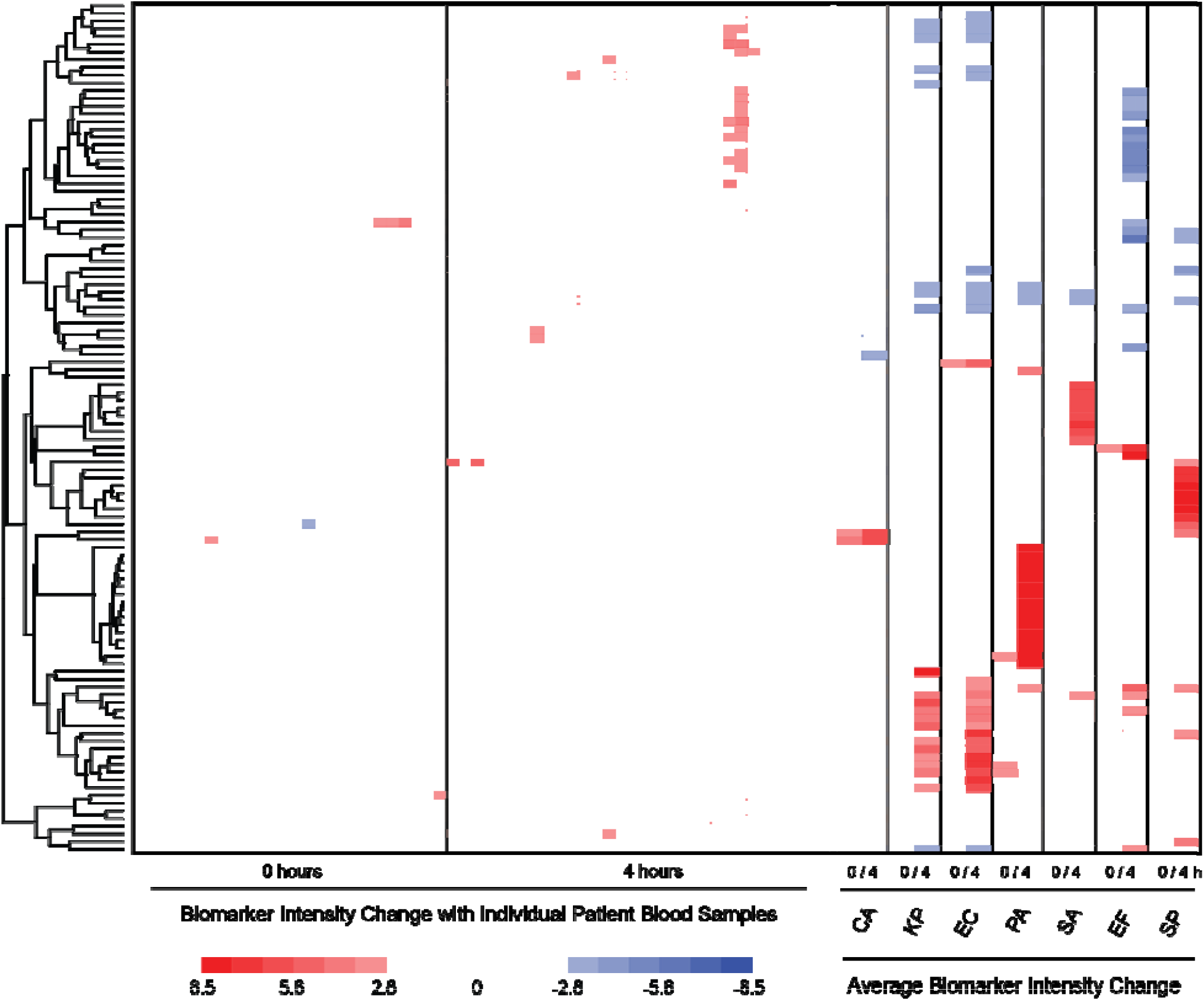
Effect of individual patient blood samples on biomarker metabolism. Individual patient blood samples (10% v:v) were incubated in Mueller Hinton medium to assess blood carryover and metabolism of top 104 biomarkers. Changes in selected biomarker intensities was negligible when compared to average changes in biomarkers when compared to Mueller Hinton medium containing 10% blood seeded with respective pathogens. See Extended Data File 4 for raw data.

**Extended Data File 1**.

**Untargeted MAVEN analysis of biomarkers at 0 and 4 hours**. Total peak intensities for each sample are shown for each m/z and retention time (RT).

**Extended Data File 2**.

**Filtered biomarkers at 4 hours (1758 to 799 peaks)**. Significant biomarkers from the initial MAVEN untargeted set were first computed by analysis of variance using a Bonferroni corrected α = 0.05. The resulting 1758 markers were then filtered to ensure that the average signal intensity for at least one organism was >20,000, and that a 2-fold change in average signal intensity compared to the Mueller Hinton Blood (MHB) medium was observed for at least one organism.

**Extended Data File 3**.

**Parent ions of significant biomarkers (104 peaks)**. Peaks were clustered into 104 groups based on retention times, potential adduct/fragment masses, and covariance of signal intensities among all replicates (equally weighted) using R software, and the parent ion, representing a real biological metabolite, was identified from each group based on highest signal intensity. Compound identifications were verified by standard additions and MS/MS (Data S4). Compounds IDs in grey have not yet been confirmed.

**Extended Data File 4**.

**Compound identification via standard additions and MS/MS**. Standards were spiked into samples at concentrations ranging from 1-1000 µM. High energy collision dissociation patterns (using collision energies from 10-50 eV) we used to confirm correct isomers.

**Extended Data File 5**.

**Blood biomarker data from 20 different patients**.

**Extended Data File 6**.

**Antibiotic susceptibility testing biomarker data**.

## Notes

### Competing Interest Statement

Drs. Lewis and Dr. Rydzak are authors on a patent relating to the use of LC-MS for detecting blood stream infections. No other competing interests declared.

### Author Declarations

Approved by the University of Calgary's Conjoint Health Research Ethics Board under certificate # REB17-1525.

