## Supplementary figures and images for "Metabolic Preference Assay for Rapid Diagnosis of Bloodstream Infections"

### Extended Data Fig. 1

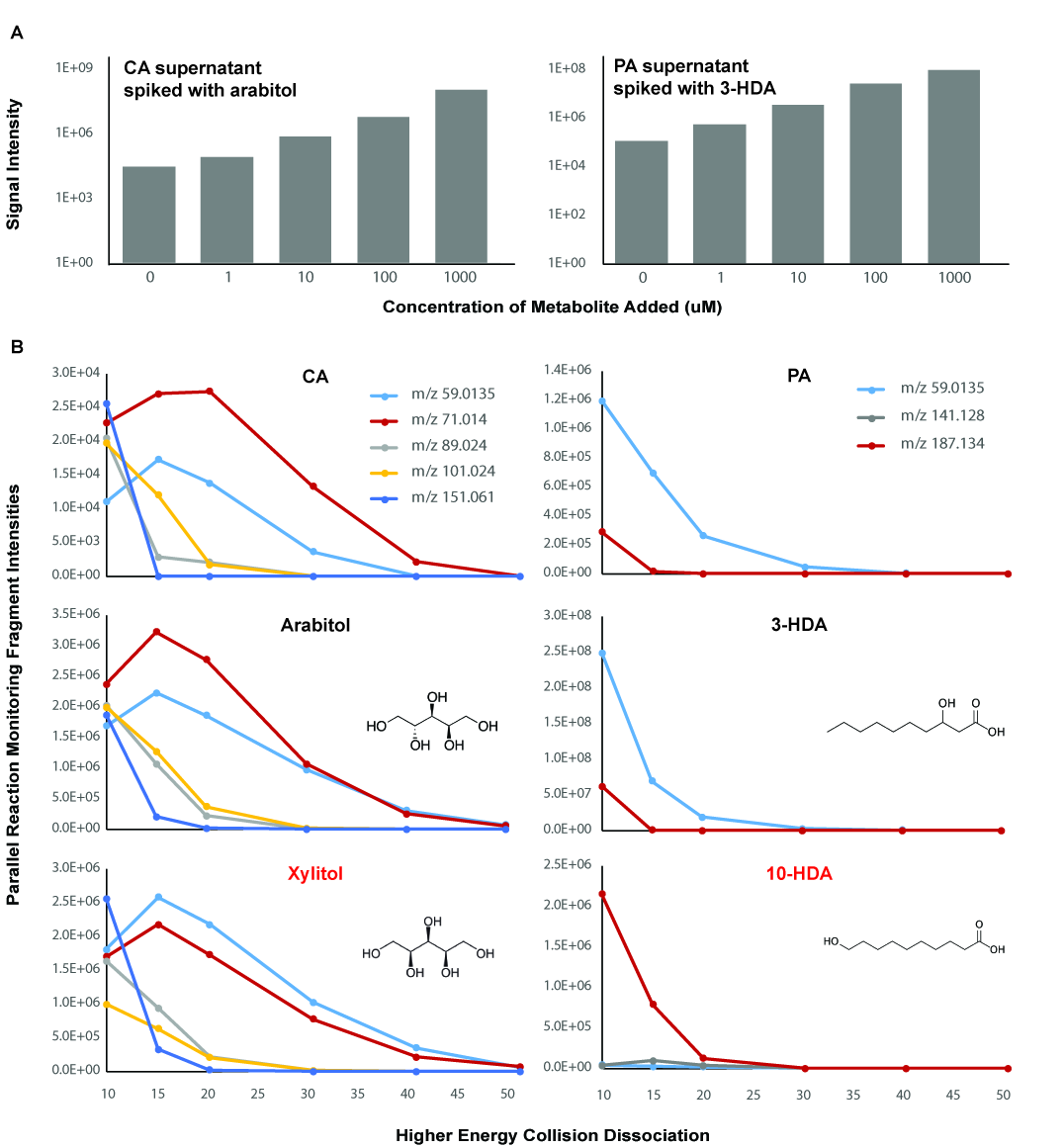

### Extended Data Fig. 2

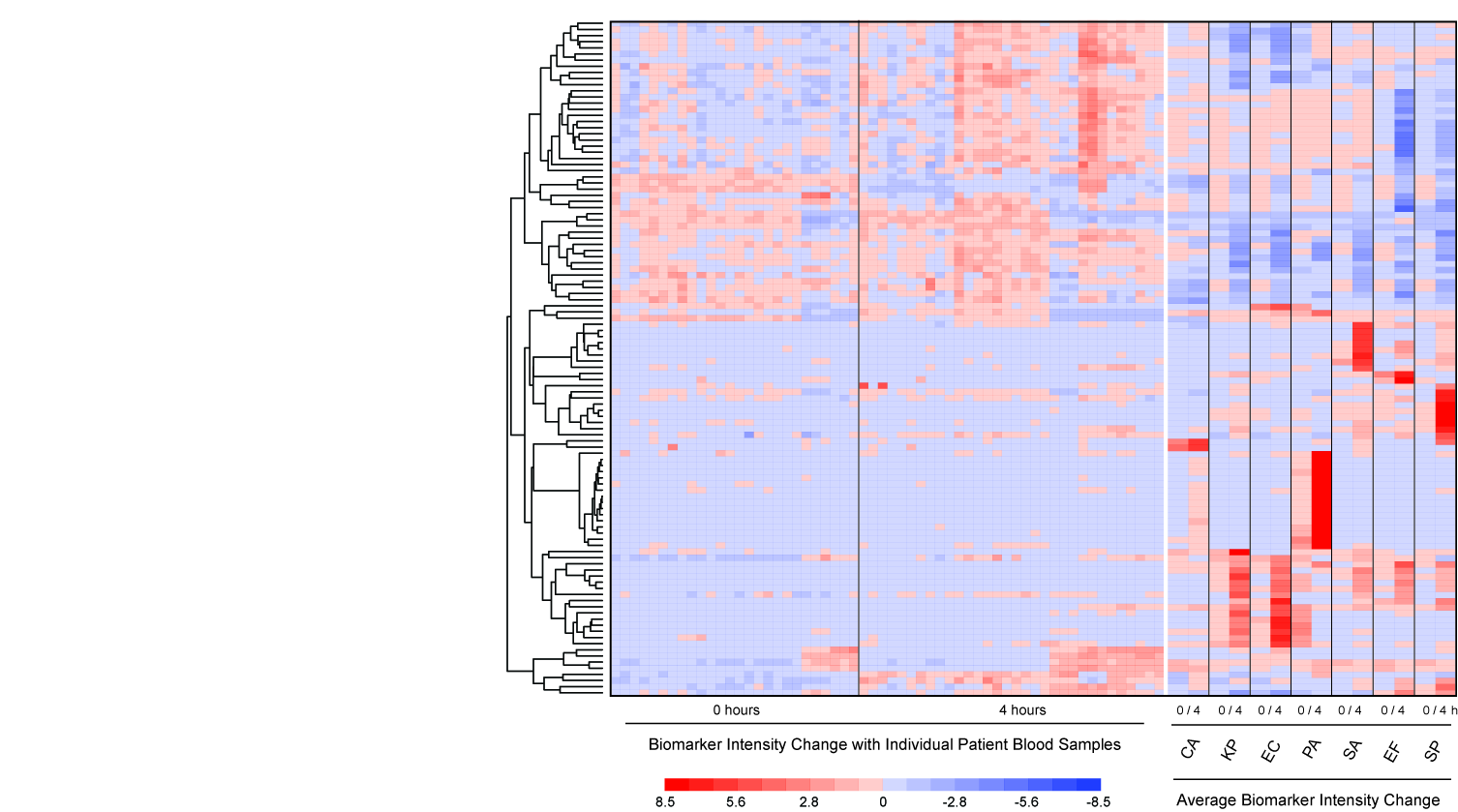
